# The impact of thrombosis and bleeding among patients with Myeloproliferative Neoplasm: Systematic Review and Meta-Analysis

**DOI:** 10.1101/2020.08.27.20182535

**Authors:** Tsegahun Worku Brhanie, Carmen Fava, Aleksandar Jovanovski

## Abstract

**Background:** Thrombosis and bleeding are the most common complications which contribute to significant morbidity and mortality of myeloproliferative patients. This study aimed to find out the incidence of thrombotic and bleeding events during diagnosis and follow up among patients with myeloproliferative neoplasm. This might help in the early detection of thrombosis and bleeding and prevention of such complications for MPN patients.

**Methods:** A systematic review and meta-analysis was conducted to assess the incidence of thrombosis and bleeding. Data extracted from the literatures in Google scholars, Mendeley, PubMed, and EMBASE databases. Studies that had thrombosis and/or bleeding reports with any types of myeloproliferative neoplasm were included in this study. We used random effect model to estimate the odd ratio, relative risk and risk difference with 95%CI of each studies and the pooled results based on Cochrane methods of Revman. A funnel plot and I^2^ test checked to see the publication bias and heterogeneity respectively.

**Results:** Nineteen studies with 14706 participants that had fitted the inclusion criteria were included in the overall thrombosis study. Five studies (n=931) included for incidence thrombosis at diagnosis and follow up. The pooled overall frequency thrombosis was 18.6%. The pooled incidence of thrombosis at diagnosis was 26.5% and odds ratio (OR= 3.17,95%CI 0.96 to10.43); relative risk (RR= 2.07,95%CI 0.98 to 4.34); risk difference (RD=21%, 95%CI −0.05 to 0.48, high certainty). Thrombosis had significant differences during diagnosis and follow up. A history of thrombosis, age >60years, and smoking were some of the risk factors for thrombosis.

**Conclusions:** Based on the findings, thrombosis and bleeding are the highest complications occurred among myeloproliferative neoplasm patients. This problem is also common both during diagnosis and follow up of MPN patients. Early detection and follow up is needed to prevent MPN complications.

## 1. Introduction

Myeloproliferative disorders are a group of hematologic malignancies in which the bone marrow makes too many red blood cells, white blood cells, and platelets. They comprise several clonal hematologic diseases that arise from a transformation in a hematopoietic stem cell. myeloproliferative neoplasm is complicated and transformed into myelofibrosis or leukemia^13^. The three main Philadelphia negative myeloproliferative disorders are polycythaemia Vera, essential thrombocythemia, and idiopathic myelofibrosis which are characterized by various combinations of erythrocytosis, leucocytosis and thrombocytosis ^1^.

The World Health Organization classification (WHO 2008) includes systemic macrocytosis, chronic eosinophilic leukaemia, chronic myelomonocytic leukaemia, and chronic neutrophilic leukaemia, polycythaemia Vera, essential thrombocythemia^2, 3^.

Polycythaemia Vera is a disease in which too many red blood cells are made in the bone marrow. Whereas primary myelofibrosis from abnormal blood cells and fibbers build up inside the bone marrow. Essential thrombocythemia causes an abnormal increase in the number of platelets made in the blood and bone marrow. Symptoms-Headache, Burning or tingling in the hands or feet, Redness, and warmth of the hands or feet, vision, or hearing problems^4, 11, 12^.

The annual incidences of both polycythaemia Vera and essential thrombocythemia are 1 to 3 cases per 100,000 population; myelofibrosis is less common^5^.

An activating mutation of Janus kinase 2 (JAK2; JAK2V617F) is present in almost all patients with polycythaemia Vera (PV), 30% to 50% of patients with essential thrombocythemia (ET) and primary myelofibrosis (PMF)^6^.

The underlying causes of myeloproliferative neoplasms are largely unknown. A mutation in the gene for Janus kinase 2, (JAK2)V617F, is present in most erythropoietin-independent erythroid colonies in polycythaemia Vera. The mutation is present in 95% of polycythaemia Vera patients and in approximately 50% of essential thrombocythemia and primary myelofibrosis patients ^7^

The increase of the thrombotic risk observed at progressively higher haematocrit values parallels blood viscosity, although also biochemical changes in the cell membrane and content could contribute to rheological abnormalities and this may lead to myeloproliferative neoplasm patient’s complication and morbidity and mortality^8^.

Therapy in both PV and ET is targeted to prevent thrombohemorrhagic Complications. On the other hand, thrombosis, leucocytosis, and age are the risk factors for the survival of patients with essential thrombocythemia and polycythaemia vera^9^.

Thrombosis and bleeding are the most common complications which contribute to significant morbidity and mortality for myeloproliferative patients^10^.

Arterial and the venous are the systems for myeloproliferative neoplasm complications to thrombosis. The Objective of this study is to find out the frequency of thrombotic and bleeding events for Myeloproliferative neoplasm patients. Early detection and prevention of thrombosis and bleeding complication is important to reduce mortality of patients with MPN.

## 2. Methods and Materials

Literature was searched by using Google scholars, Mendeley, PubMed, and EMBASE databases for studies on thrombosis and myeloproliferative neoplasm. Terms used to search were “thrombosis, bleeding and myeloproliferative neoplasm”. By using the terms, we got a total of 3144 articles; in Google scholar 2340 articles, in PubMed 287 articles, in Mendeley 190 articles and in EMBASE 327 articles (recorded as #1). Finally, thirty-two articles were selected and included in this study, which were fitted the inclusion criteria.

We filtered human studies with the English language and sorted by best match and recent studies. A Systematic Review and Meta-analysis were done based on the PRISMA guideline. A Citation and references were managed by Mendeley desktop. All types of studies having thrombosis and myeloproliferative neoplasm and their clinical characteristics. We compared studies, patients with myeloproliferative neoplasm with thrombosis, and without thrombosis. Age and sex differences of thrombosis also considered. We used relative risk (RR) to measure the effect size difference and confidence interval for each study. We also used a random effect to measure the relative weight assigned to each study. Publication bias was checked by using a funnel plot.

Heterogeneity between studies was calculated using the I^2^test.

### The inclusion criteria

Patients who were diagnosed MPN (either PV, ET and PMF or all types)

Studies that have information about thrombosis, and/or bleeding in patients with myeloproliferative neoplasm, studies that have information about the study participants’ age, and sex. Whereas studies that have not full information for our study, or studies that did not report thrombosis, and/or bleeding among myeloproliferative disorder were excluded.

## 3. RESULTS

Data extraction and the selection of literature were conducted as shown by the following diagram.

**Table 1.**
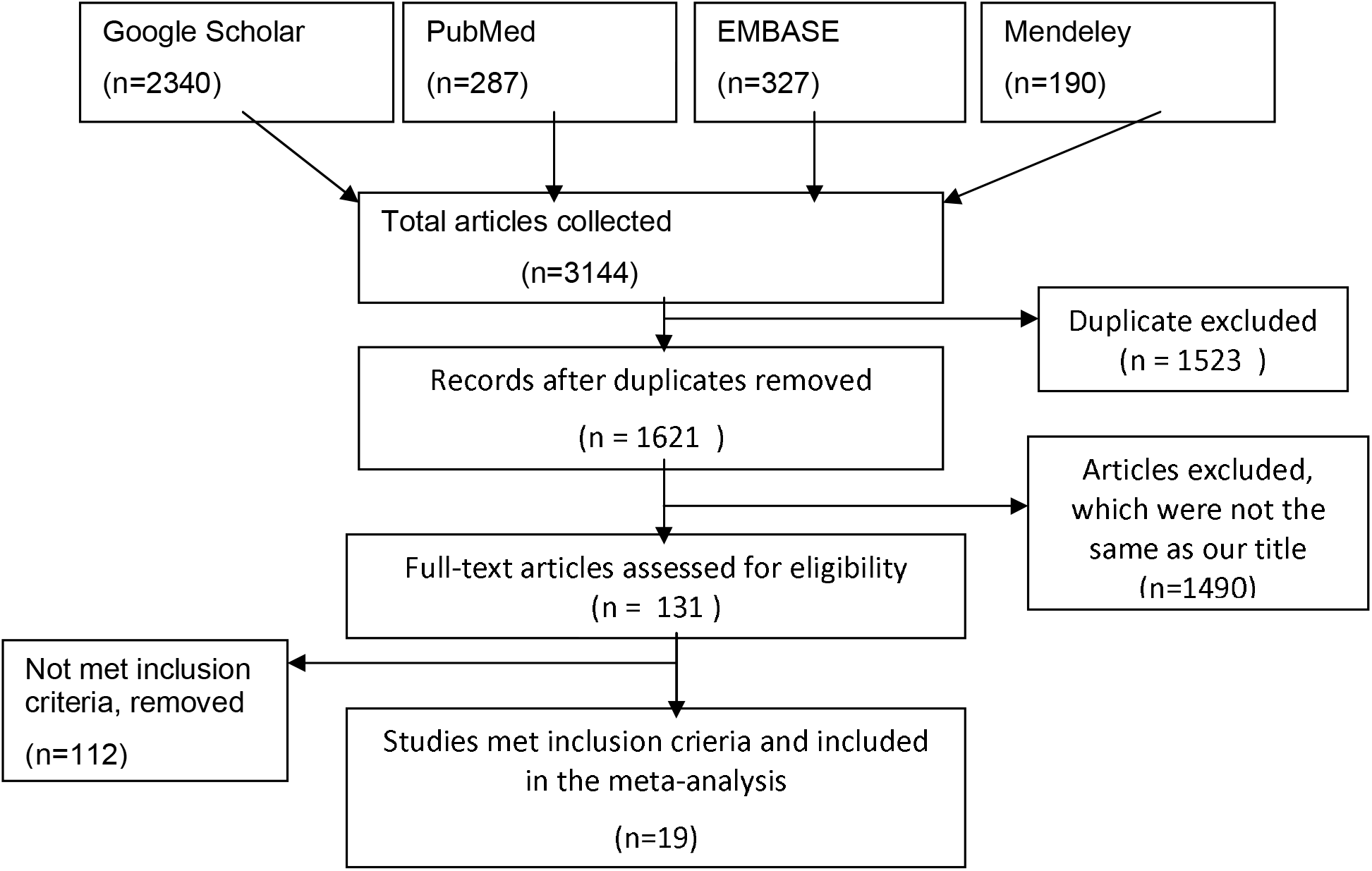
The general characteristics of included studies

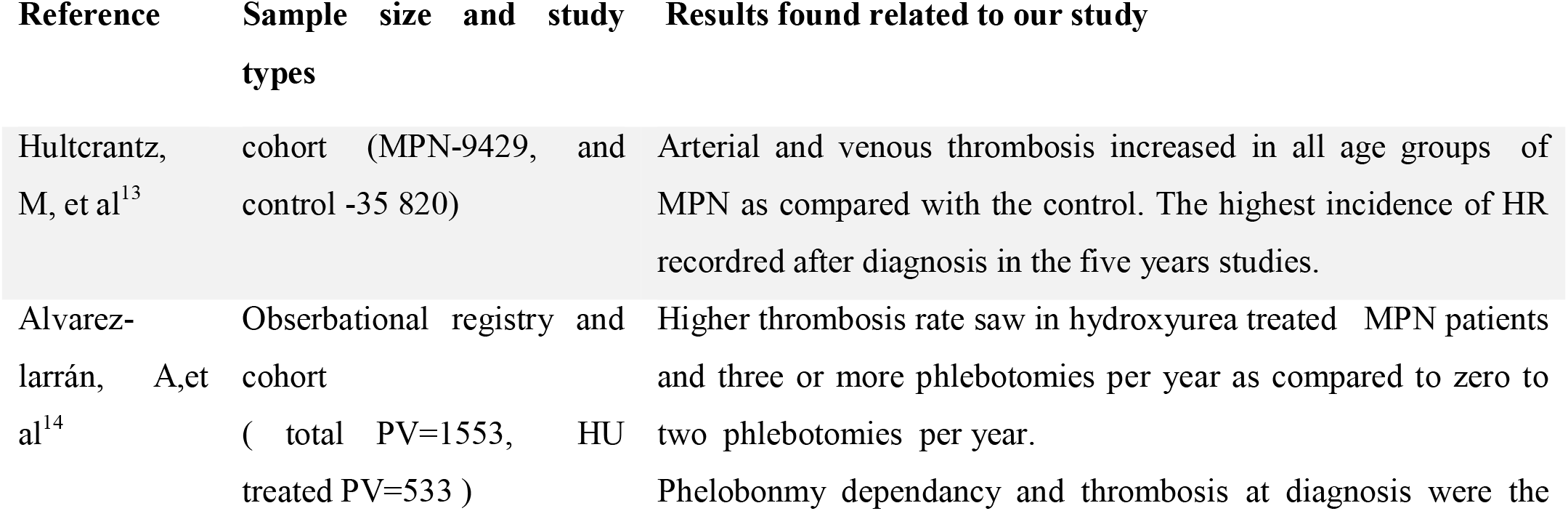
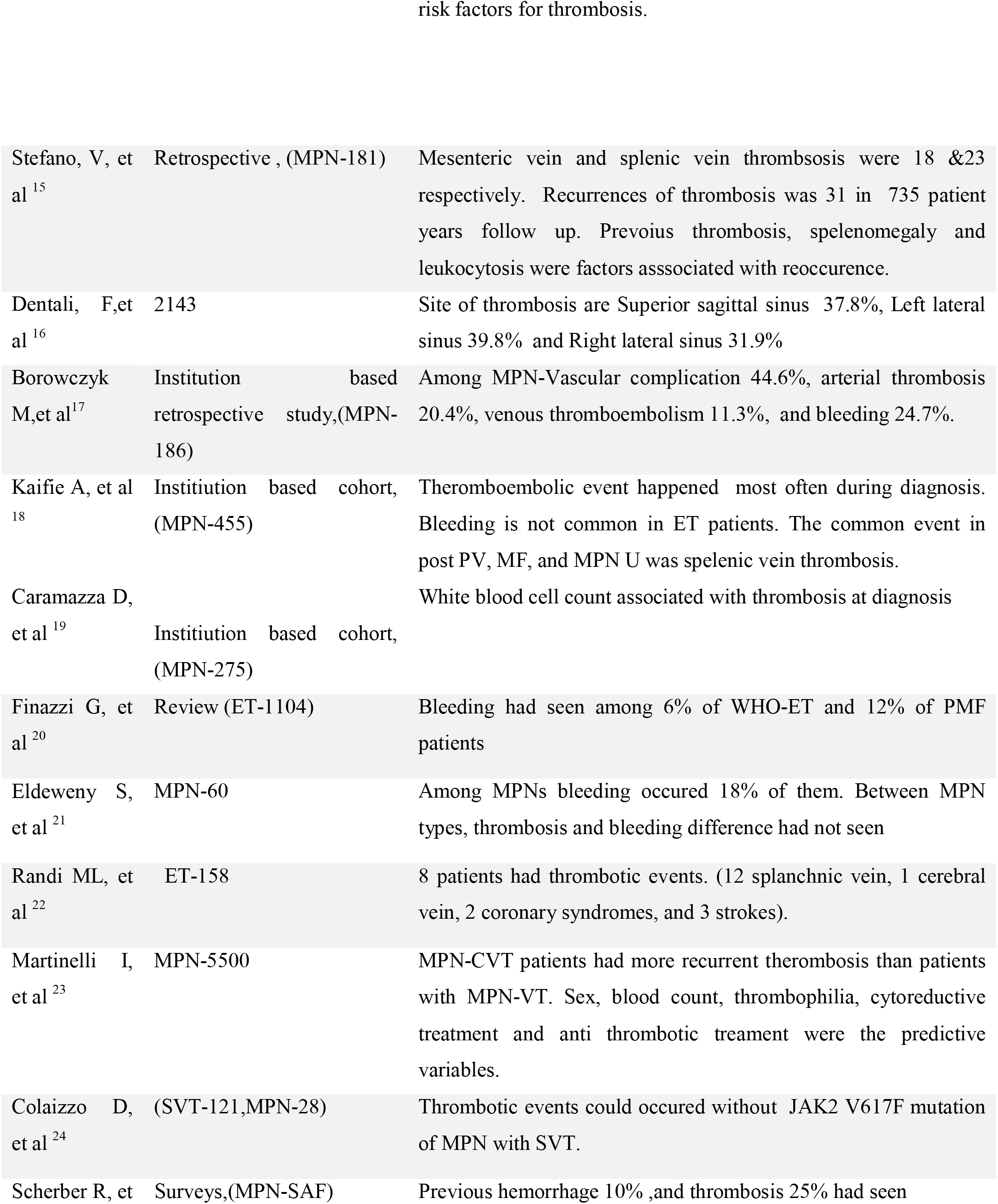
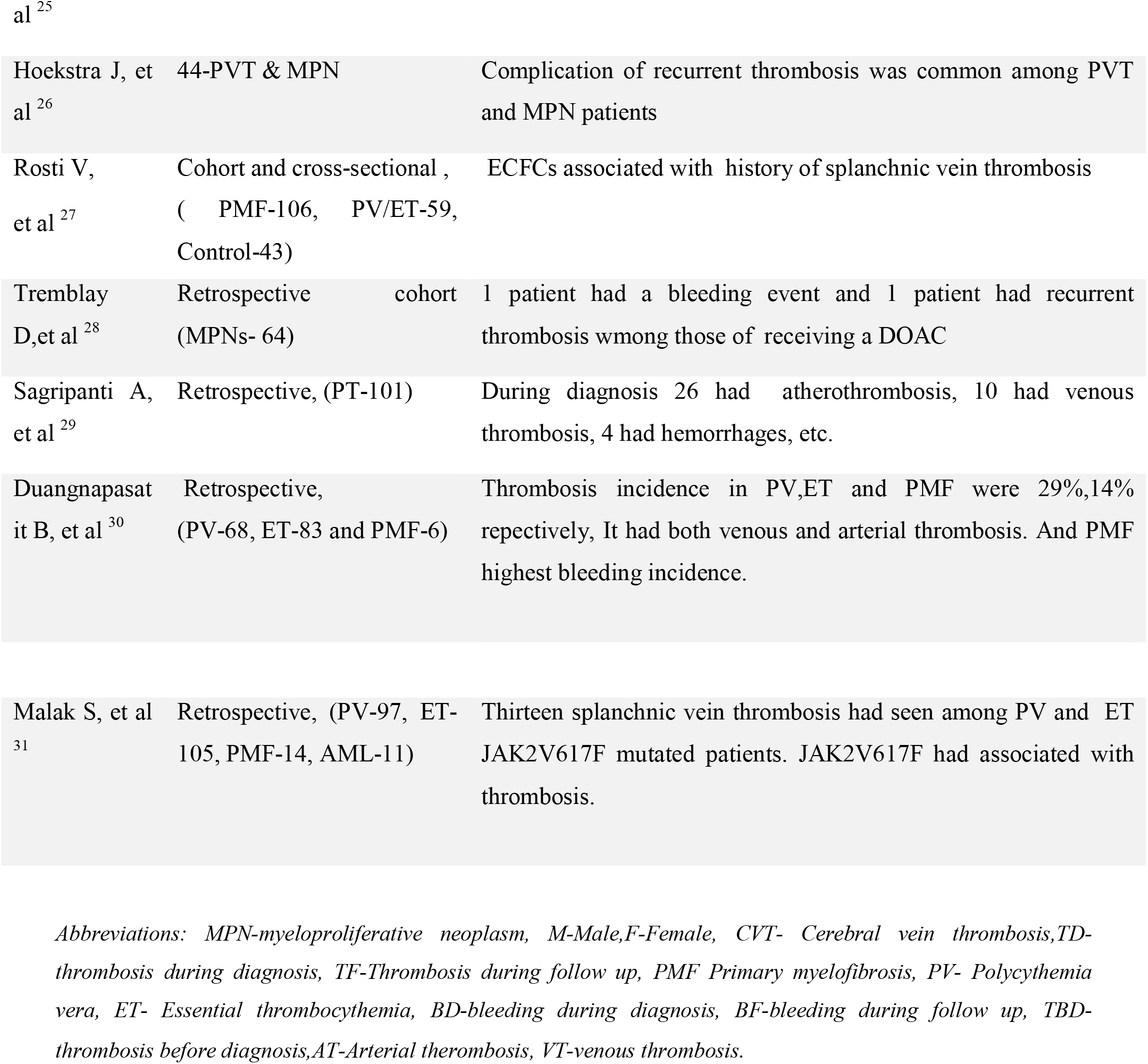

The pooled result of meta-analysis showed that, the odd of developing thrombosis among MPN patients was 17% and it had statistically significant (P=0.0008) in random effect model. And thrombosis incidence during diagnosis and follow up had not statistically significant difference (P=0.06). see it the forest plot fig 1 and fig 2 below.

**Fig 1.**
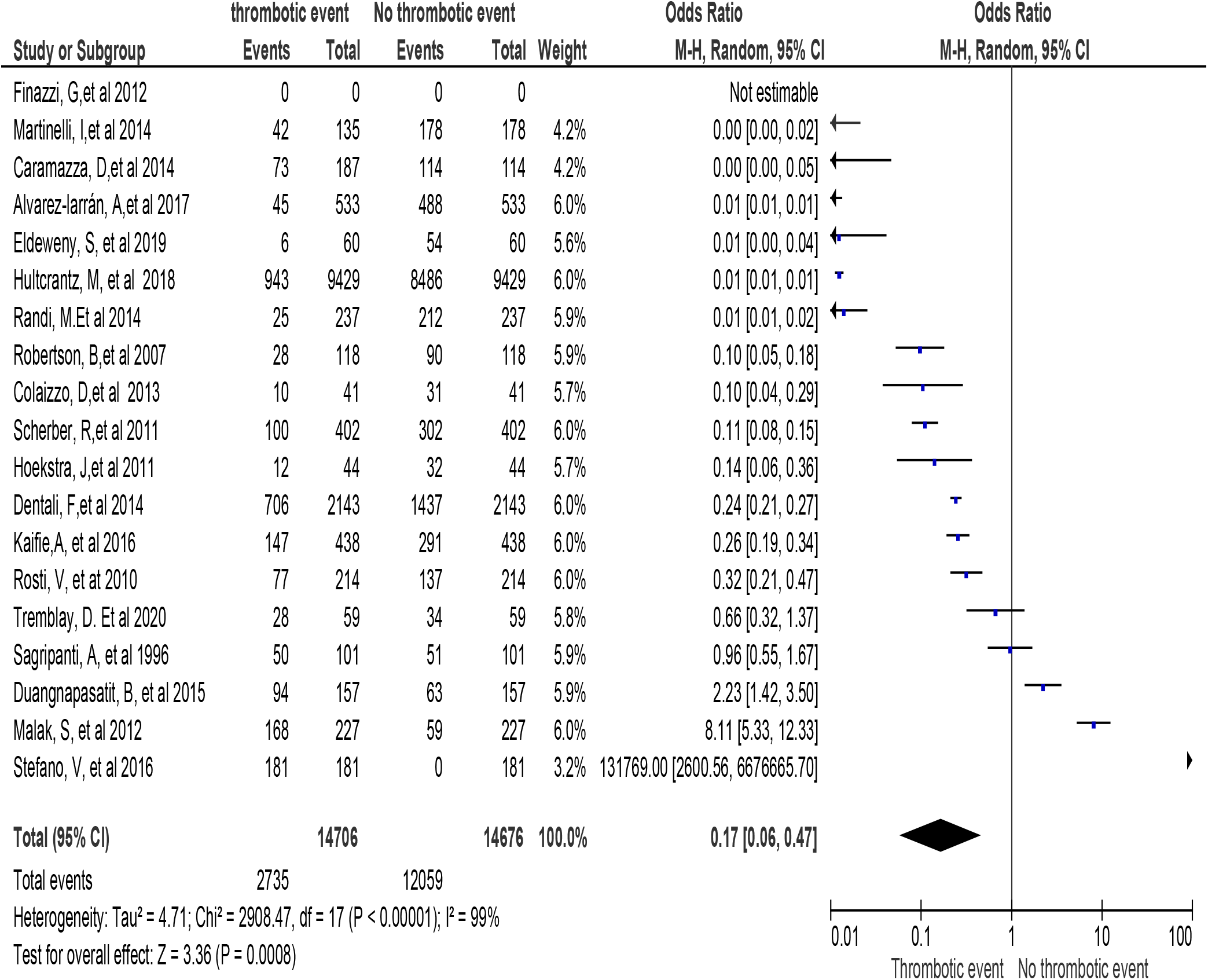
Forest plot for pooled prevalence of thrombosis among patients with MPN.

**Fig 2.**
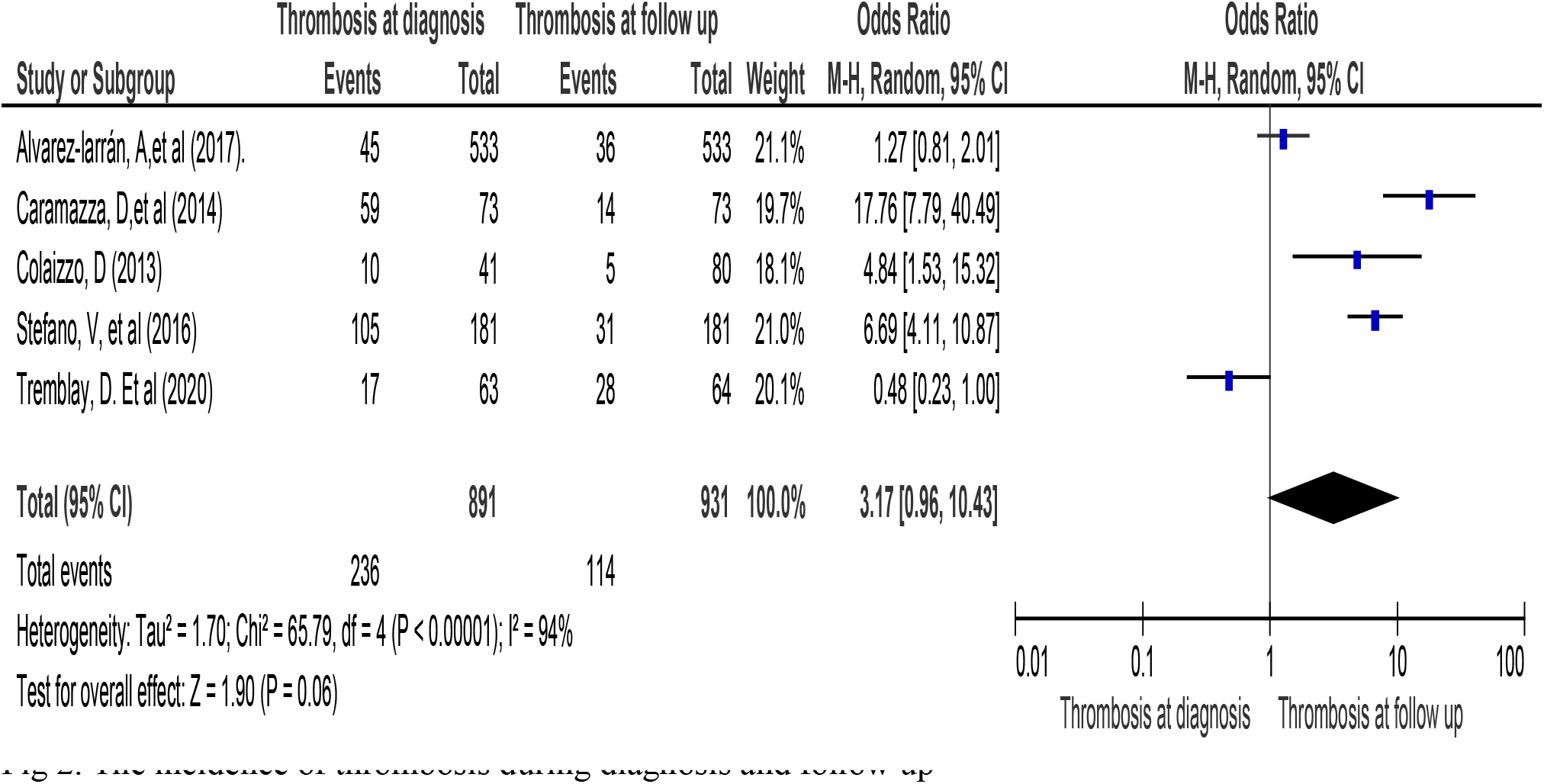
The incidence of thrombosis during diagnosis and follow up

When we compare thrombosis events between PV and ET patients, it had statistically significant difference (P=0.03). that mean, thrombotic events were higher in PV than in ET. see it the next figure 3.

**Figure 3.**
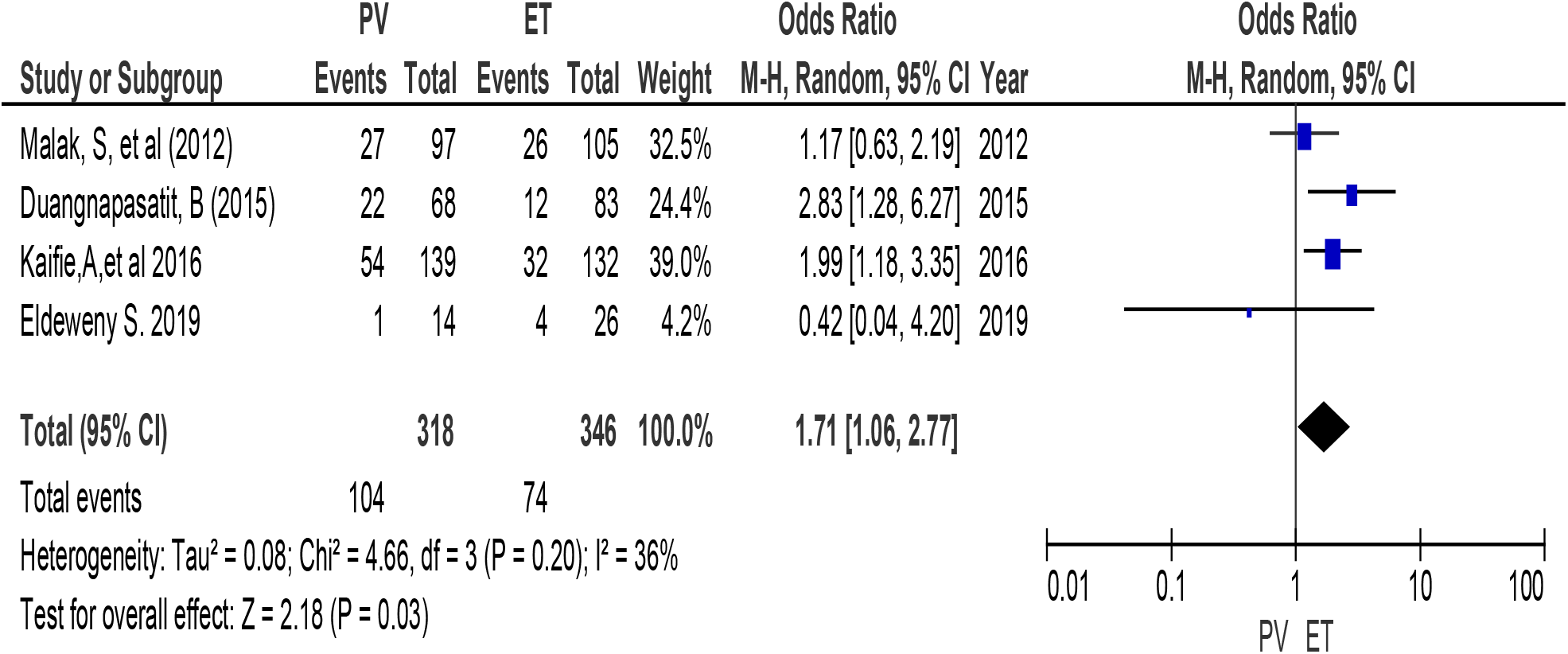
Forest plot for thrombosis incidence between PV and ET

The risk difference of thrombosis between PV and ET patient had not statistically significant (P=0.011), look figure 4 below.

**Figure 4.**
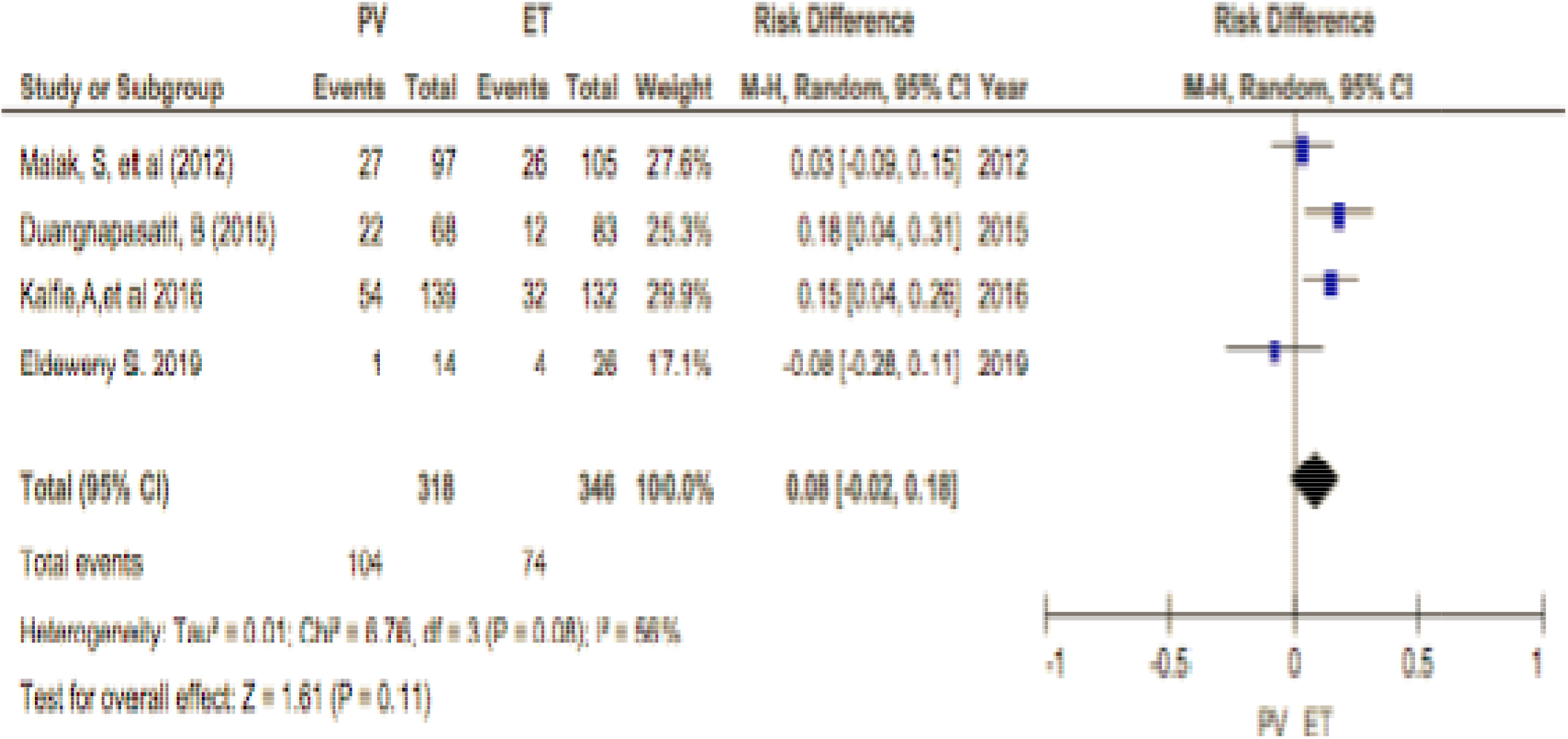
Forest plot for thrombosis risk difference between PV and ET patients.

**Figure 5.**
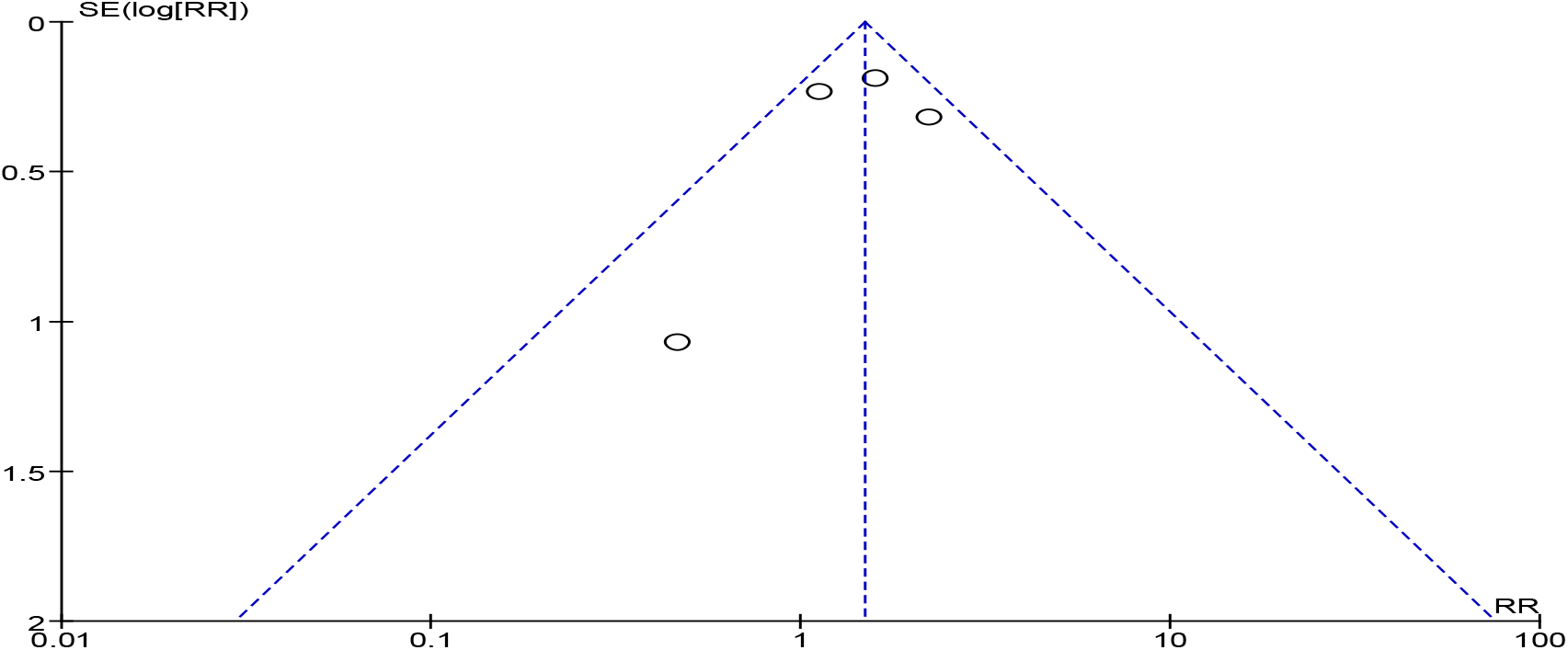
Funnel plot

Bleeding prevalence between PV and ET patients was not statistically significant, (P=0.098). See fig 6 below.

**Figure 6.**
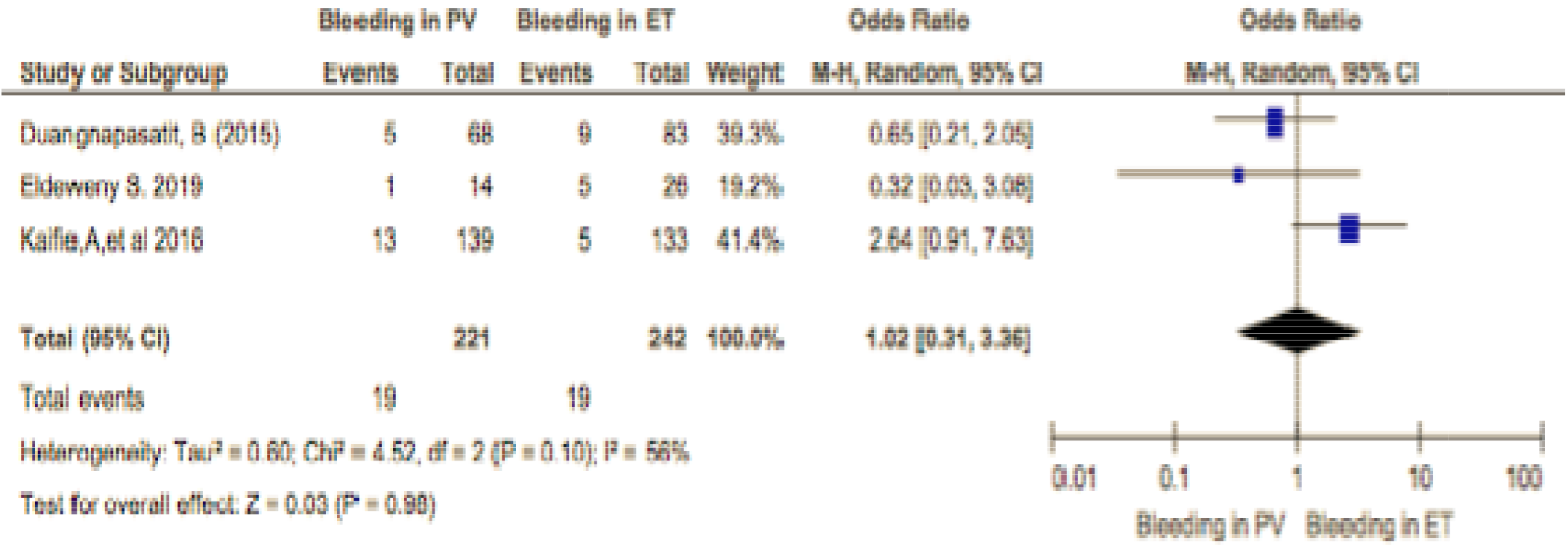
Forest plot for pooled bleeding prevalence among patients with PV and ET.

The risk difference of bleeding between PV and ET patient had not statistically significant (P=0.93), look figure 7 below.

**Figure 7.**
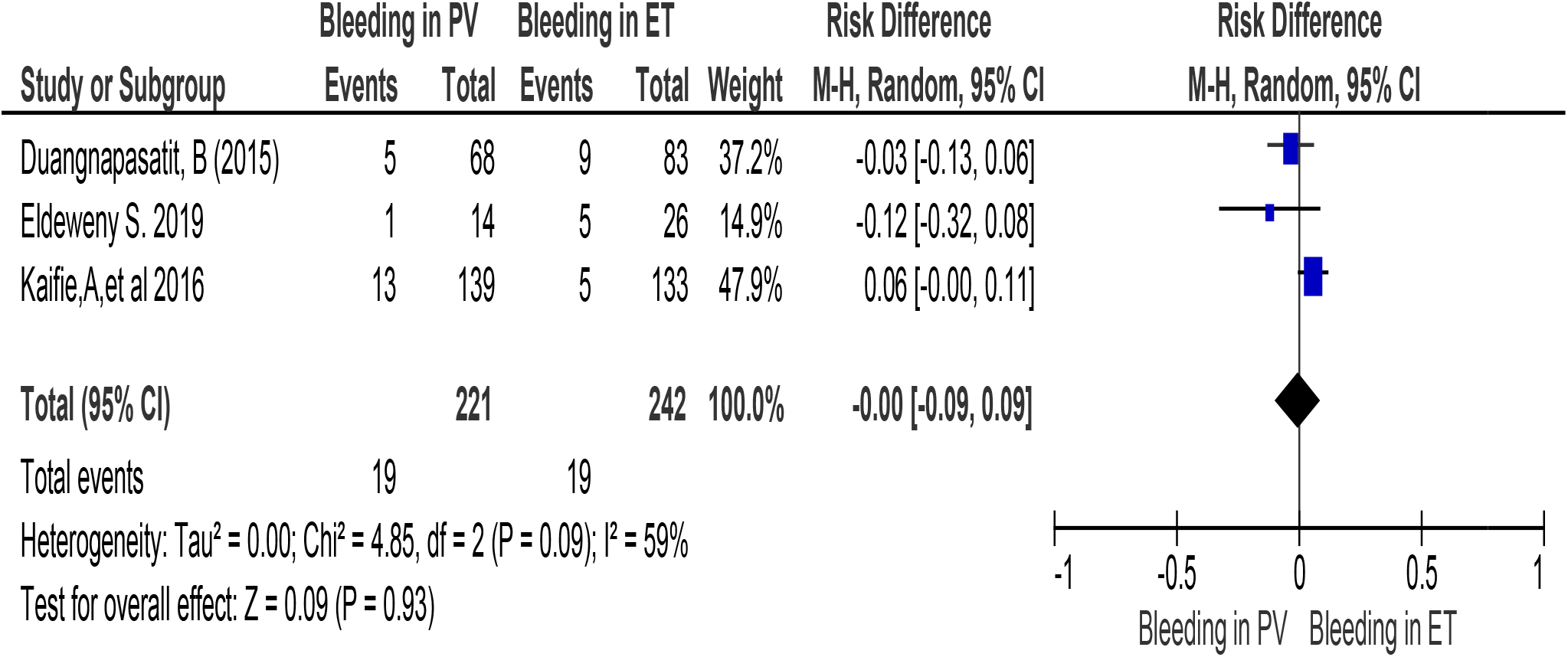
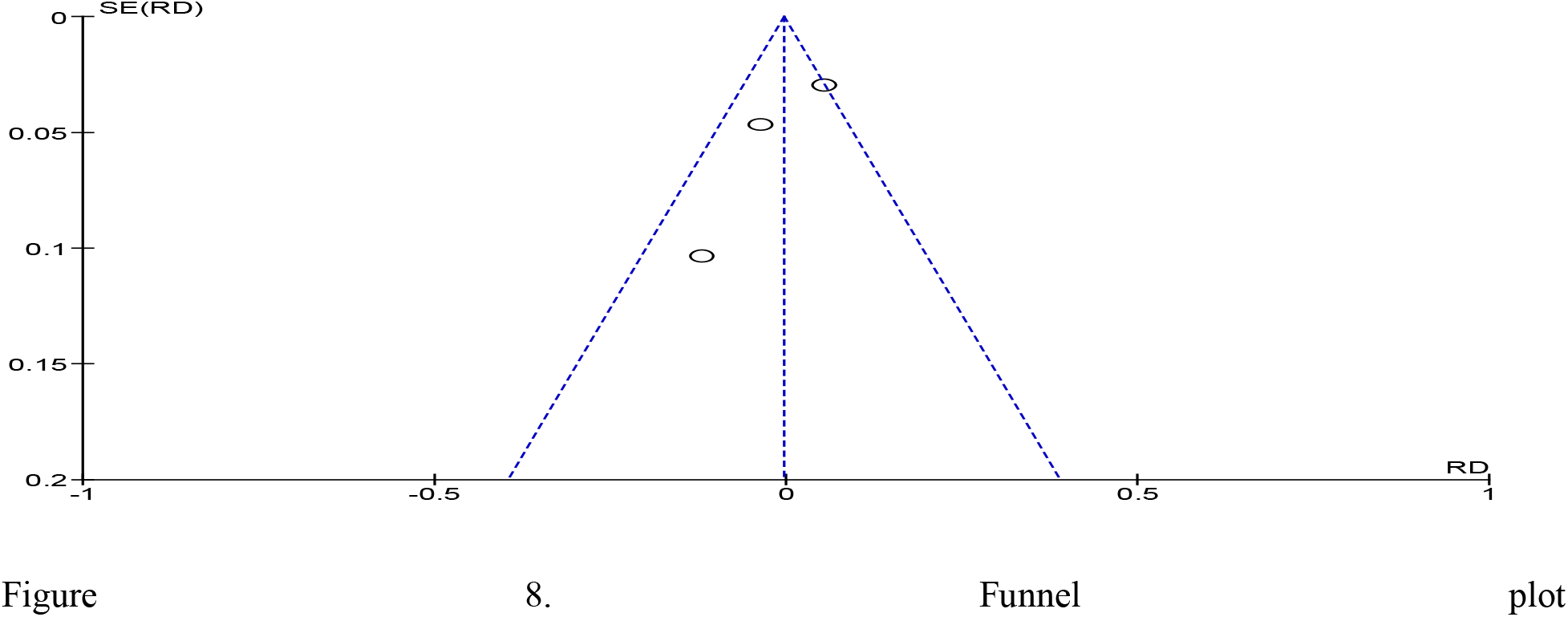
Forest plot for Bleeding risk difference between PV and ET patients.

## 4. DISCUSSION

Thrombosis and bleeding are the most common complication that cause the morbidity and mortality of patients with myeloproliferative neoplasm^14,16,19,21,22,23,29,29,37,38,38,40,41^. This review showed that among a total of 14706 MPN patients, 2735 thrombotic events happened and there are also many bleeding recorded. Venous and arterial thrombosis has been the highest prevalence site of events. There are many contributing factors for such complication. Age, previous thrombosis, late diagnosis and follow up are some of them^17, 32^. They have different site of thrombosis and the site oi bleeding.

More than 9.5×10^9^_L_ white blood cells which has lowered by hydroxyurea therapy is associated with thrombosis^19^. Among MPN patients, the incidence of thrombosis was high in PV and ET patients. It had both arterial and venous event. Whereas the incidence of bleeding was high in patients with PMF. Thrombosis and bleeding had been the highesl burden recorded in Ph negative MPN patients^30^.

Mesenteric vein and splenic venous thrombosis found to be greater and BCS, previous thrombosis history, splenomegaly, and leukocytosis are the associated factors. Extracranial and intracranial bleeding have the major one^15^. The PV and ET therapies use to lower thrombosis complication. Age greater than 60 years and previous thrombosis are vascular risk predictors ^33^.

Bleeding occur during follow up among patients with ET and PMF and, diagnosis of PMF, leukocytosis, aspirin therapy and past history of hemorrhage are bleeding predictors^20^.

Ninety two percent of the investigated cases have vascular risk factors and the complication occur within 12 months after ET and PV diagnosis^34^. The incidence of arterial and venous thrombosis is increasing across in all age groups of MPN patients in comparison with the controls. The thrombosis rate is highest after diagnosis and then it decreases there after^13^.

The prevalence of CALR mutations is lower among MPN, BCS and PVT patients, and thrombosis rate is lower in CALR Mutation as compared with JAK2V617F mutation^35^.

The mortality rate of MPN patients with JAk2V617F mutation are higher. The length of KAK2V617F mutation development is 21 months during the follow up. Thrombosis free survival is smaller and there is no bleeding incidence difference between groups^24^.

The thrombosis of splenic vein is correlated with MPN, eighty percent of thrombosis occurred among patients with MPN. Esophageal variceal bleeding recorded due to portal hypertension^28^.

Among a study of 44 PVT and MPN patients, the median age during diagnosis was forty-eight years and most of the patients were females. Eleven percent and twenty three percent of the patients had previous thrombosis and family history of thrombosis respectively. Many factors that causes venous thrombosis in addition to MPN^26^. Thrombosis phenotypes are different across MPN types in mesenteric and vein thrombosis ^36^.

14 myocardial infractions found in 263 patients. Ninety three percent of patients have vascular risk factors. MI have seen after 12 months of ET or PV diagnosis. Platelet activation and leukocytes role in MPN associated thrombosis helps for thrombosis prevention and estimation^34^.

**Limitation of the study:** different study designs had included in this study, which may reduce strength of the evidence. Literature search were limited with English language, this also reduce the access of studies in other language. And unpublished information had not included in this study.

## 5. CONCLUSIONS

Bleeding and thrombosis have the common complication faced among Myeloproliferative neoplasm patients. The incidence of thrombosis is the major problem both during diagnosis and follow up. Age greater than 60 years, previous thrombosis, leukocytosis and other factors are associated with thrombosis among patients MPN.

## Data Availability

Data is not available

## Acknowledgement

I would like to thank you partnership for Knowledge and Torino University for their scholarship opportunity to my study.

## Contributions

TWB-literature search, analysis, wrote and edited the manuscript.

## CF-comments and correction

AJ-idea and suggestion

